# Blood metabolomic shift links diet and gut microbiota to multiple health outcomes among Hispanic/Latino immigrants in the U.S.

**DOI:** 10.1101/2024.07.19.24310722

**Authors:** Yang Li, Brandilyn A. Peters, Bing Yu, Krista M. Perreira, Martha Daviglus, Queenie Chan, Rob Knight, Eric Boerwinkle, Carmen R. Isasi, Robert Burk, Robert Kaplan, Tao Wang, Qibin Qi

**Affiliations:** Department of Epidemiology and Population Health, Albert Einstein College of Medicine, Bronx, NY 10461, USA; Epidemiology, Human Genetics & Environmental Sciences, School of Public Health, University of Texas Health Science Center at Houston, Houston, TX 77225, USA; Department of Social Medicine, University of North Carolina at Chapel Hill, Chapel Hill, NC 27599, USA; Institute for Minority Health Research, University of Illinois at Chicago, Chicago, IL 60612, USA; Department of Epidemiology and Biostatistics, School of Public Health, Imperial College London, United Kingdom; Departments of Pediatrics and Computer Science & Engineering, University of California San Diego, La Jolla, CA 92093-0763, USA; Human Genetics Center, University of Texas School of Public Health, University of Texas Health Science Center at Houston, Houston, TX 77225, USA

**Author notes:** **Corresponding author** Qibin Qi, Department of Epidemiology and Population Health, Albert Einstein College of Medicine, Bronx, NY 10461, USA.

**Keywords:** Blood metabolome, U.S. lifestyle, acculturation, gut microbiota, cardiometabolic traits, obesity, diabetes, chronic kidney disease

## Abstract

Immigrants from less industrialized countries who are living in the U.S. often bear an elevated risk of multiple disease due to the adoption of a U.S. lifestyle. Blood metabolome holds valuable information on environmental exposure and the pathogenesis of chronic diseases, offering insights into the link between environmental factors and disease burden. Analyzing 634 serum metabolites from 7,114 Hispanics (1,141 U.S.-born, 5,973 foreign-born) in the Hispanic Community Health Study/Study of Latinos (HCHS/SOL), we identified profound blood metabolic shift during acculturation. Machine learning highlighted the prominent role of non-genetic factors, especially food and gut microbiota, in these changes. Immigration-related metabolites correlated with plant-based foods and beneficial gut bacteria for foreign-born Hispanics, and with meat-based or processed food and unfavorable gut bacteria for U.S.-born Hispanics. Cardiometabolic traits, liver, and kidney function exhibited a link with immigration-related metabolic changes, which were also linked to increased risk of diabetes, severe obesity, chronic kidney disease, and asthma.

**Graphical abstract:** 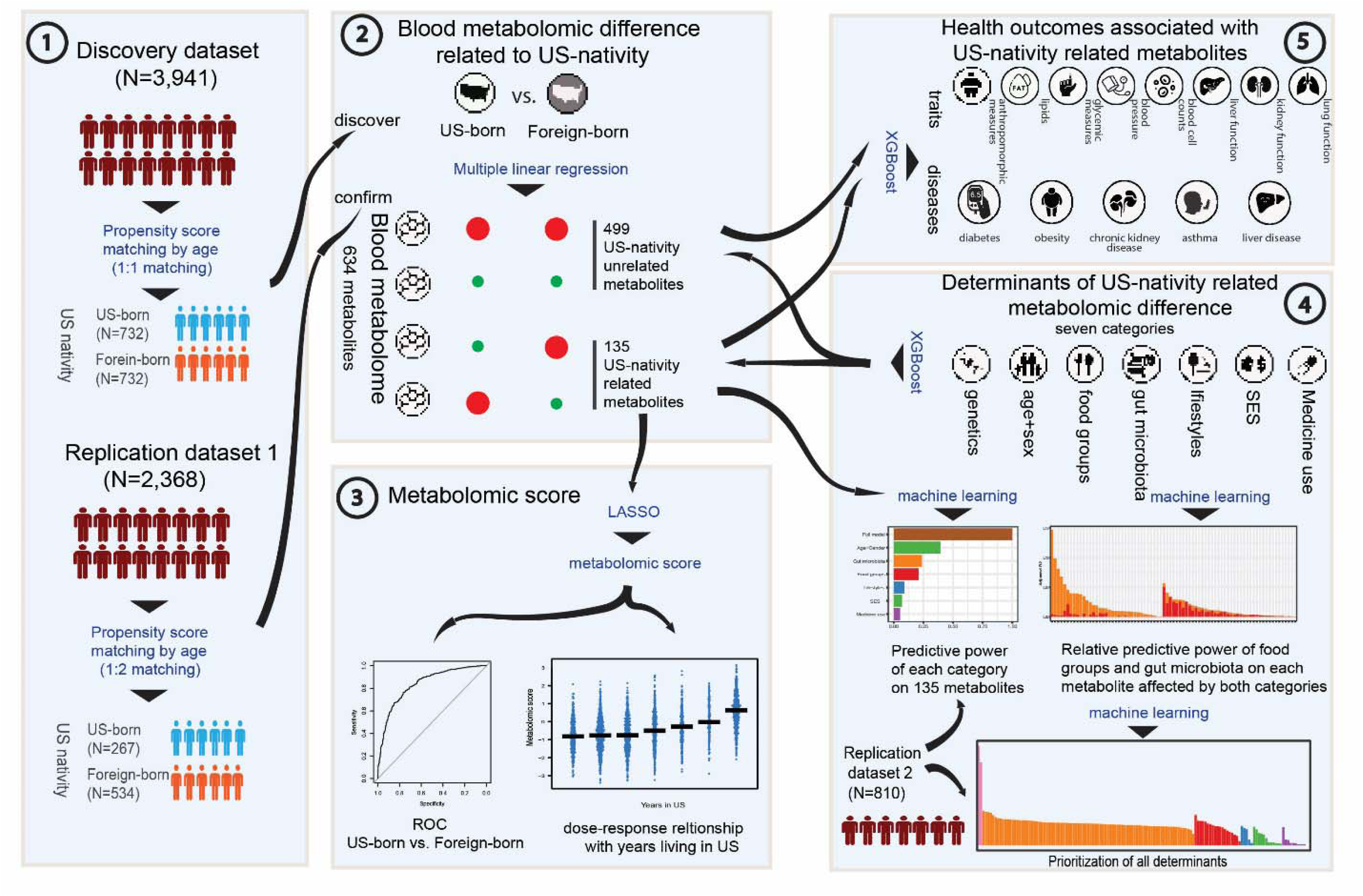

**Highlights:**

- A substantial proportion of identified blood metabolites differ between U.S.-born and foreign-born Hispanics/Latinos in the U.S.
- Food and gut microbiota are the major modifiable contributors to blood metabolomic difference between U.S.-born and foreign-born Hispanics/Latinos.
- U.S. nativity related metabolites collectively correlate with a spectrum of clinical traits and chronic diseases.

## Introduction

Immigrants to the U.S. from less industrialized countries or U.S. territories face an elevated risk of obesity, diabetes, liver diseases and other chronic diseases compared to individuals of the same ethnicity who continue to reside in their countries of birth [1–4]. Additionally, longer residence in the U.S. is associated with worse health condition. This health disparity might be predominantly attributed to adoption of the U.S. lifestyle habit, which is a feature of acculturation [5]. After relocating to the U.S. 50 states/DC, immigrants often experience substantial shifts in food habits, lifestyles, socioeconomic status, access to medical care, and overall environment [5]. However, a comprehensive examination to determine the exact risk factors contributing to this elevated chronic disease burden during their transition to life in the U.S. is yet to be fully conducted.

The blood metabolome (i.e. the complete set of small molecules in blood) may represent a unique tool to understand the effects of U.S. acculturation on health. The blood metabolome contains rich systemic information crucial for understanding physiology and predicting various chronic illnesses [6]. In addition, blood metabolites are influenced by a multitude of environmental and genetic factors [7–9]. Among these, diet and gut microbiota have been noted as the main sources of blood metabolites [7, 8, 10–12]. For U.S. immigrants, the shift towards U.S. lifestyles often entails diets enriched in processed foods, added sugars and unhealthy fats, with reduced intake of nutrient-dense whole foods [13–15]. U.S. acculturation can also shape gut microbiota, leading to reduced microbiome diversity and function [16]. Other factors that can impact blood metabolites, like lifestyle factors [7, 17–19], socio-economic status (SES) [20] and drug intake [21] are subjected to change after arriving to the U.S. [1, 22, 23]. Therefore, examining the interplay between blood metabolome, various factors, and health outcomes may help advance our understanding of elevated disease risk among immigrants during the acculturation.

Hispanics/Latinos are the largest minority population in the U.S. [24]. By examining differences in the blood metabolome between U.S.-born Hispanics/Latinos and foreign-born Hispanics/Latinos, and the determinants of those differences, we can potentially uncover valuable insights into the contribution of acculturation to immigrant health. Here, we used machine learning to scrutinize the blood metabolome-mediated link between lifestyle risk factors and multiple health outcomes, among 1,141 U.S.-born (i.e., born in U.S. 50 states/D.C.) and 5,973 foreign-born Hispanics/Latinos from the Hispanic Community Health Study/Study of Latinos (HCHS/SOL).

## Methods

### Study cohort

The HCHS/SOL is a prospective study of Hispanic/Latino populations in the U.S. From 2008 to 2011, 16,415 adults aged 18–74 years, self-identified as having Cuban, Dominican, Puerto Rican, Mexican, Central American or South American heritage, were recruited from a random sample of households in four communities (Bronx, NY; Chicago, IL; Miami, FL; and San Diego, CA). 2,863 participants were born in the 50 U.S. states, and 13,479 were born in the nations of Latin America or the territories of the U.S. Participants were recruited by using a 2-stage probability sample design, and data in the current study was collected at the baseline visit and six-year follow-up visit. The study protocol was approved by the institutional review boards of all collaborating institutions, and written informed consent was obtained for all participants.

Based on the visit when blood samples were collected and the batch when blood metabolome was profiled, the data for the entire cohort were divided into three datasets: discovery dataset (N=3,941. Blood samples from baseline visit; metabolome was profiled at the first batch), replication dataset 1 (N=2,368. Blood samples from baseline visit; metabolome was profiled at the second batch), and replication dataset 2 (N=810. Blood samples from follow-up visit; metabolome was profiled at the second batch). This division aimed to address the potential batch effects in metabolomic profiling caused by profiling batch and this visit. More details about blood sample collection and data splitting are included in Supplemental methods.

Discovery dataset and replication dataset 1 served to identify and confirm U.S. nativity related blood metabolites. To minimize the well-known impact of age on blood metabolome, we conducted age matching at 1:1 ratio within discovery dataset and 1:2 ratio within replication dataset 1, according to the proportion of U.S.-born and foreign-born participants. The procedure aimed to create sub-datasets for specific analyses. Replication dataset 2 specially served to compare the association of different types of determents with blood metabolites, as blood sample from metabolome profiling and stool sample for metagenome were collected at the same visit.

### Metabolomic profiling of blood samples and Metagenomic profiling of stool samples

Blood metabolomic profiling was performed using an untargeted liquid chromatography-mass spectrometry (LC-MS) based protocol on discovery HD4 platform at Metabolon (Durham, North Carolina, USA). Participants were asked to fast for ≥8 h before the examination. Detailed procedures have been described elsewhere [25] and Supplemental methods. Metabolomic data was normalized using Quantile Normalization. After removing unknown metabolites and the metabolites which were undetected in greater than 20% samples, 634 metabolites were finally obtained for the following analyses in the current study.

Metagenomics Sequencing was performed on DNA extracted from fecal samples collected by FTA card using a novel shallow-coverage method of shotgun sequencing-based Illumina NovaSeq platforms. Microbiome bioinformatics analyses, taxonomic assignment, and functional components identification were performed using the SHOGUN pipeline as described previously [26] and in Supplemental methods. Metagenomic data was normalized and transformed using Centered Log-Ratio method (CRL). After removing these genus which were prevalent in less than 20% samples or whose average read counts across samples were less than 10, 87 genus were included in the following analyses.

### Identification of US-related metabolites and derivation of metabolomic score

Multiple linear regression model with each metabolite as dependent variable was used to identify different metabolites between U.S.-born participants and foreign-born participants while controlling for age, sex, visit center, Hispanic background, and five principal components of population structure. These metabolites which were identified in the matched discovery dataset (P<0.05 after False Discovery Rate [FDR] correction) and confirmed in the matched replication dataset 1 (P<0.05) were defined as U.S. nativity related metabolites. The rest metabolites were considered as U.S. nativity unrelated metabolites.

Least Absolute Shrinkage and Selection Operator (LASSO) was applied to predict U.S. nativity status (U.S.-born as 1 and foreign-born as 0) with U.S. nativity related metabolites using glmnet package, from which the predictive value was generated and used as metabolomic score. The training and testing of LASSO model were performed in different datasets. The predictive performance of metabolomic score in discriminating U.S.-born and foreign-born participant was measured by calculating Area Under the Receiver Operating Characteristic (AUROC).

### Association between determinant categories and metabolites

The potential determinants of blood metabolites were classified into seven categories, including genetics, age+sex, food groups (19 food groups), gut microbiota (87 microbes at the genus level), lifestyle (factors related to smoking, drinking, physical activity, sedentary time, and sleep duration), socioeconomic status (SES), and medication use (20 drugs). Explained variance (EV) was calculated to measure the association between each metabolite and each determinant category using eXtreme Gradient Boosting (XGBoost) implemented in xgboost package. All XGBoost models were trained using 80% samples in the dataset and testing in 20% samples. EV from testing dataset were used for the following statistical analysis and visualization. Adjusted EV (EV_adj_) was additionally calculated for food groups and gut microbiota. The detailed determinants in each category and algorithm of EV calculation were described in Supplemental methods.

For genetics, we calculated SNP-based heritability of metabolites, instead of EV, to measure its association with each metabolite. Genotyping and imputation protocols and heritability estimation have been described previously [27] and in Supplementary methods.

To measure the feature importance of each determinant and find specific associations between features and metabolite levels, Shapley additive explanations (SHAP) values were calculated based a full model including all non-genetic determinants using SHAPforxgboost package as described in Supplement methods.

### Predictive power of determinants on overall U.S. nativity related metabolites

Predictive power on each metabolite was estimated for each determinant category using XGBoost, except genetic factors which used heritability instead of predictive power. To estimate the predictive power of different determinant category (separated model) and all determinants (full model), we first applied PCA over all U.S. nativity related metabolites data to get the first 100 PCs which constitute more than 98% of the total variance in the data. XGBoost was used to predict the PCs on the basis of the determinant categories independently. EV of each PCs were calculated separately and then combined as predictive power for each determinant category:

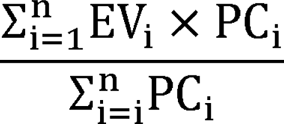

where EV_i_ is the EV that XGBoost recovered for the i^th^ PC. PC_i_ is the fraction of variance that the i^th^ PC explains out of the overall variation of 100 PCs. n is the total number of PCs.

A full model which included all non-genetic determinants was built to predictive power for all non-genetic determinants. Relative predictive power was the percentage of predictive power of each category model in the full-model predictive power.

### Association between metabolites and clinical traits

The investigated clinical traits include continuous variables from cardiometabolic traits, chronic inflammation marker, blood cell counts, liver function indices, kidney function indices, and lung function indices, as described in Supplemental methods. The EV of each clinical trait was calculated using XGBoost with U.S. nativity related metabolites or U.S. nativity unrelated metabolites as the estimate of association between metabolites and clinical traits. All traits were standardized before entering the model. To compare the predictive performance of two group metabolites while controlling the bias caused by the number of input variables, Bootstrap sampling was applied to obtain 1000 metabolite set with 25 metabolites from two groups, respectively. Thus, 1000 EV values for U.S. nativity metabolites and 1000 EV for U.S. nativity unrelated metabolites were finally harvested to compare the predictive performance using permutation test for each clinical trait.

### Association between metabolites and disease outcomes

Five representative diseases were investigated in the study, including type 2 diabetes (T2D), obesity, chronic kidney disease (CKD), asthma, and Metabolic dysfunction-associated fatty liver disease (MAFLD) as described in Supplemental methods. The principle of Gene Set Enrichment Analysis (GSEA) was used to test the enrichment of U.S. nativity related metabolites in all metabolites which were ranked by their association with diseases to reflection the overall association between U.S. nativity related metabolites and a certain disease. Firstly, Poisson regression model was fitted with the incidence of disease as output and individual metabolite as input with adjustment for age, sex, visit center, Hispanic background, and the first 5 PCs of population structure. As the metabolite level was normalized and comparable, the beta values of metabolite were used to rank all metabolites in the descending order. Next, the ranked metabolite list and 135 U.S. nativity related metabolites were used as the input for the MSEA algorithm implemented by fgsea package. Normalized enrichment score and P-value were given for U.S. nativity related metabolites with each disease. P-value was determined by permutation testing, comparing the observed enrichment score to scores obtained from randomly permuting the gene labels. The results of MESA were visualized using enrichment plots, showing the enrichment score and leading-edge subset of genes contributing to the enrichment signal.

In addition to MSEA, Poisson regression model was fitted between disease outcomes and metabolomic score with adjustment for age, sex, visit center, Hispanic background, and the first 5 PCs of population structure to measure the association (represented by RR and 95% CI) between each U.S. nativity related metabolite and the incidence of each disease outcome.

### Quantification and Statistical Analysis

R 4.1.2. and related packages were used for all statistical analysis, machine learning, and visualization of results. Age matching was performed using MatchIt 4.4.0. Multiple regression model and LASSO regression were carried out using base 4.1.2 and glmnet 4.1.4, respectively. pROC 1.18.0 was used to calculate the discriminative power of metabolomic score. SNP-based heritability and EV were calculated using GENESIS 2.24.2 and xgboost 1.6.0.1, respectively. SHAP feature importance was extracted from XGBoost model with SHAPforxgboost 0.1.1. MSEA algorithm was implemented in fgsea 1.20.0. ggplot2 3.3.6 and ComplexHeatmap 2.10.0. were used for visualization. Adjusted and raw P values <0.05 were considered to be significant.

## Results

### Overview of the study Cohorts

In the discovery dataset (N=3,941), mean age was 36.5 years among 732 U.S.-born participants (53.1% females), and mean age was 48.0 years among 3,209 foreign-born participants (58.1% females) (**Table S1**). In the replication dataset 1 (N=2,368), mean age was 44.9 years and 53.5 years, and 59.6% and 65.2% were females for U.S.-born (N=267) and foreign-born participants (N=2,101), respectively (**Table S1**). After being matched by age to minimize its impact on identifying U.S. nativity related metabolites, there was no difference in age between in U.S.-born and foreign-born participants in both discovery (N=1,464) and replication dataset 1 (N=801) (**Table S2**). For analysis of identifying potential determinants of blood metabolites, especially food and gut microbiota, we formed replication dataset 2 which included 810 participants with a mean age of 45.9 years, and 60.5% of these participants were females (**Table S1**). Other basic characteristics for U.S.-born and foreign-born participants in unmatched and matched datasets are shown in **Table S1** and **Table S2**, respectively.

### Blood metabolomic differences between U.S.-born and foreign-born Hispanics

We employed multiple linear regression to identify metabolites associated with U.S. nativity (U.S.-born vs. foreign-born) in the age matched discovery dataset and the age-matched replication dataset 1. These analyses adjusted for multiple covariates including age, sex, study sites, Hispanic background, and the first five principal components (PCs) of continental ancestry. The results on the associations of metabolites with U.S. nativity were highly consistent between two datasets. (Pearson’s r=0.85) (**Figure 1A**). We identified 158 metabolites associated with U.S. nativity in the discovery dataset (FDR<0.05), and then confirmed 135 of them with the same directions of associations with U.S. nativity in the replication dataset 1 (raw P<0.05) (**Figure 1A**). Most of these confirmed U.S. nativity related metabolites were amino acids (44; 32%), lipids (40; 29.6%) and xenobiotics (19; 14.1%) (**Figure 1B**). Less lipids were found to be related to U.S. nativity compared to total number of profiled lipids (P<0.05) (**Figure 1B**). In contrast, amino acids, xenobiotics, nucleotides, as well as cofactors and vitamins, were more presented in U.S. nativity related metabolites compared to total profiled metabolites, although Fisher’s exact test did not show significant enrichment of U.S. nativity related metabolites in these super-pathways (**Figure 1B**). The top 50 metabolites associated with U.S. nativity are shown in **Figure 1C**, and the association for all metabolites can be found in **Table S3**. Beta-cryptoxanthin, S-methylcysteine sulfoxide, carotene diol, 3-phenylpropionate, and hydroxy-CMPF were the top five metabolites that were higher in foreign-born participants compared to U.S.-born participants, and gamma-tocopherol/beta-tocopherol, piperine, orotate, ursodeoxycholate, and 2-hydroxyoctanoate were the top five metabolites that were higher in U.S.-born participants compared to foreign-born participants.

**Figure 1.**
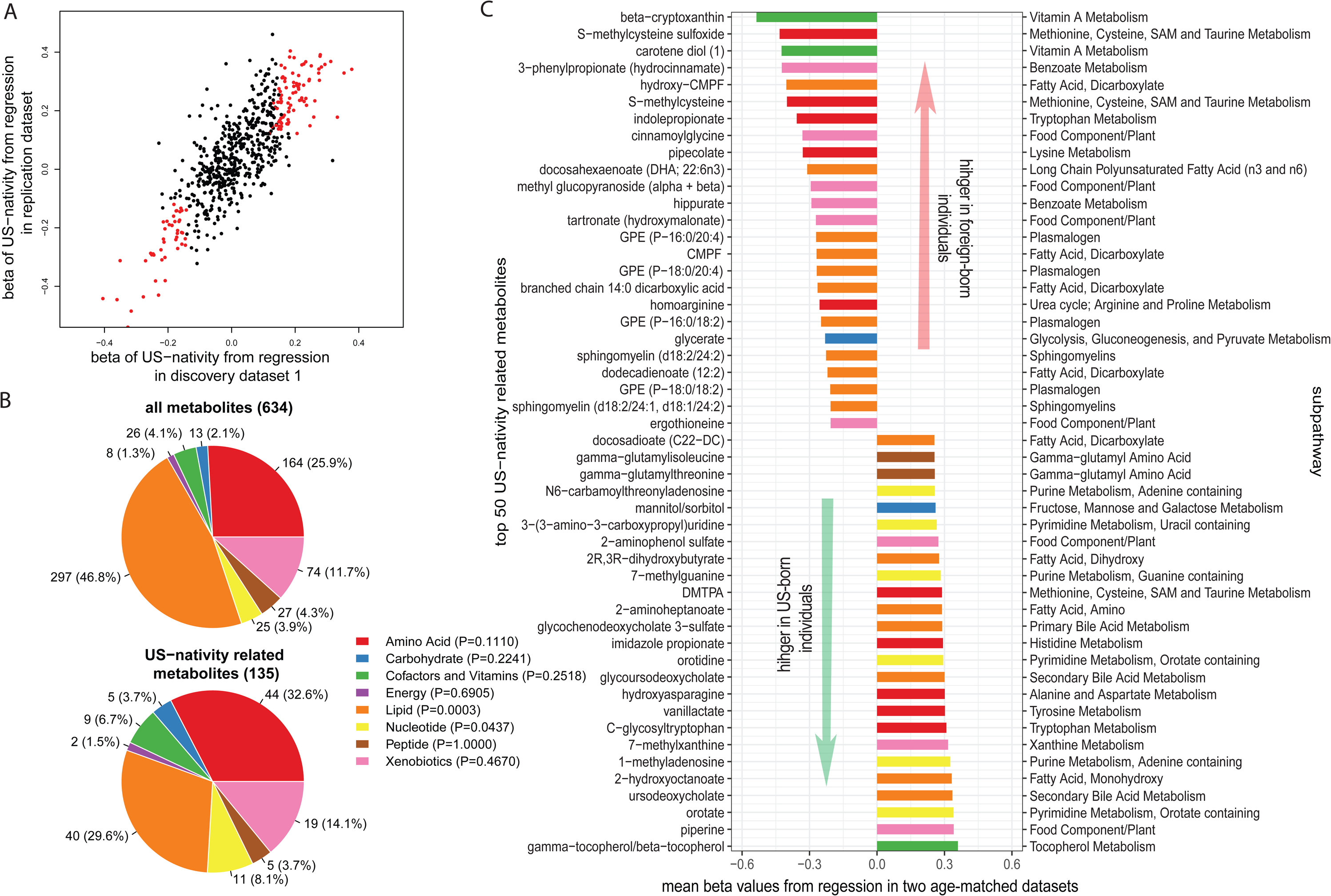
U.S. nativity related blood metabolites. (A) Consistency between U.S. nativity-blood metabolite associations from age-matched discovery dataset and their associations from age-matched replication dataset. The associations between U.S. nativity and metabolites were measured by β values from multiple linear models with adjustment for age, sex, visit center, Hispanic background, and five principal components of population structure. β values from discovery dataset and replication dataset are plotted against x-axis and y-axis, respectively. Red dots indicate 135 U.S. nativity related metabolites which were confirmed after validation with replication dataset and False Discovery Rate (FDR) adjustment for multiple tests. **(B)** Distribution of total 634 metabolites and 135 U.S. nativity related metabolites on the super-pathway level. Enrichment of U.S. nativity related metabolites in each super-pathway was marked behind the labels of super-pathway, as shown by P values. **(C)** Top 50 U.S. nativity metabolites selected by the average beta value from discovery dataset and replication dataset. Metabolites higher in the blood of foreign-born participants (negative β values) and those higher in the blood of U.S.-born participants (positive β values) are listed at the top and bottom, respectively. The color denotes the super-pathway for each metabolite, whose color corresponds to super-pathways in figure B.

### Discrimination of U.S.-born and foreign-born Hispanics by blood metabolomic profile

The next step of analysis was to define a nativity-related metabolomic score which reflected overall blood metabolomic differences between U.S.-born Hispanics (i.e., higher score) and foreign-born US Hispanics (lower score). We performed LASSO analysis to predict U.S. nativity status based on 135 U.S. nativity metabolites (**Figure 2**). The LASSO model was built in training dataset including 80% participants in the age-matched discovery dataset (N=1,172), tested in the remaining 20% participants, and further validated in the replication dataset 1 with all participants (N=2,368). The discrimination ability for U.S. nativity status was relatively high in all three sub-datasets (Area Under the Receiver Operating Characteristic (AUROC)=0.837, 0.840, and 0.823, respectively) (**Figure 2A**).

**Figure 2.**
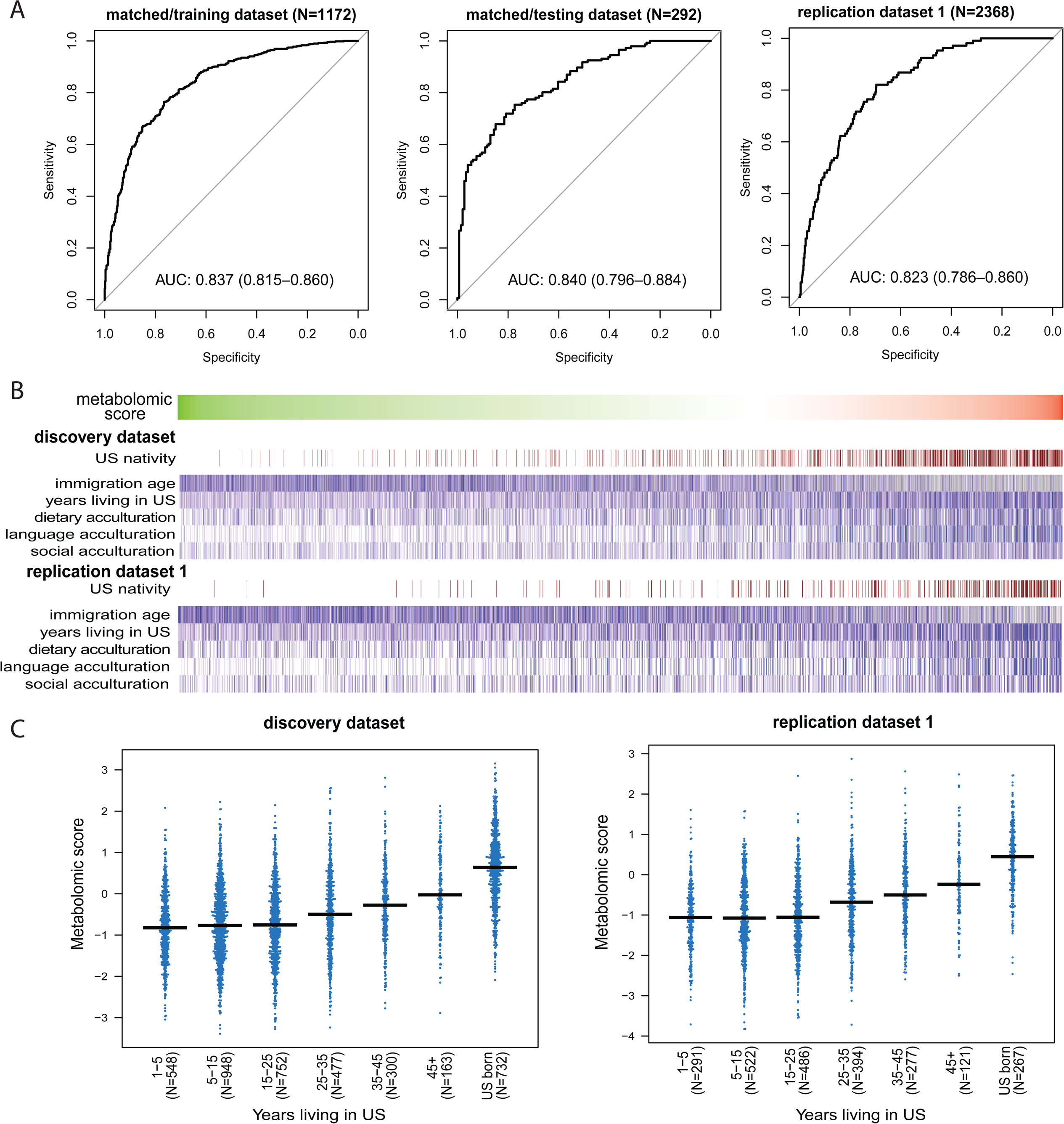
Metabolomic score associates with immigration-related variables. (A) Receiver operating characteristic (ROC) curve showing the performance of metabolomic score on predicting U.S. nativity status. Age-matched discovery dataset was divided into training dataset (60%) and testing dataset (40%). The metabolomic score was constructed with 135 U.S. nativity related metabolites using Least Absolute Shrinkage and Selection Operator (LASSO) in training dataset, validated in testing dataset, and further confirmed in replication dataset. (B) the correlation between metabolomic score and immigration-related variables, including immigration age, years lived in US, dietary acculturation score, language acculturation score, and social acculturation score. Columns are participants who are sorted by metabolomic score with low scores at the left and high scores at the right. (C) Dose-response relationship between metabolomic score and US-immigration status in discovery dataset and replication dataset 1. Foreign-born participants were divided into six groups according to years lived in the U.S., while U.S.-born participants were classified into one group. Metabolomic score for participants in each group was dotted, and the mean values were shown as bars.

We further examined associations of the metabolomic score with immigration/acculturation-related variables (**Supplemental method**) in both discovery dataset and replication dataset 1, and found that higher levels of the metabolomics score were associated with younger immigration age (Pearson’s r=-0.22, P<0.05; r=-0.23, P<0.05), longer years lived in the U.S. (Pearson’s r=0.33, P<0.05; r=0.37, P<0.05), higher dietary acculturation score (Pearson’s r=0.29, P<0.05; r=0.26, P<0.05), language acculturation score (Pearson’s r=0.42, P<0.05; r=0.38, P<0.05), and social acculturation score (Pearson’s r=0.27, P<0.05; r=0.24, P<0.05) (**Figure 2B**). We also observed a dose-response relationship between the metabolomic score and years lived in the U.S. (**Figure 2C**). Results were highly consistent between the discovery and replication dataset 1.

### Genetic and non-genetic determinants of U.S. nativity related blood metabolomic difference

To identify the potential determinants of metabolomic difference between two U.S. nativity groups, we systematically investigated SNP-based heritability (h^2^_snp_) and the predictive power of non-genetic determinants (Explained variance, EV) for each individual metabolite (**Table S4**). The working hypothesis was that genetic factor did not contribute U.S. nativity related blood metabolomic difference which were attributed to environmental factors. We first compared h^2^_snp_ and EV between 135 U.S. nativity related metabolites and the remaining 499 U.S. nativity unrelated metabolites. We did not observe significant difference in h^2^_snp_ between two metabolite sets but found two metabolite sets had different EV of non-genetic determinants in both discovery dataset and replication dataset 1 (**Figure 3A-B**). In terms of each determinant category, food groups, gut microbiota, SES, and medication use showed different EV between two metabolite sets. However, demographics and lifestyles did not show different EV between two metabolite sets in both datasets (**Figure 3A-B**).

**Figure 3.**
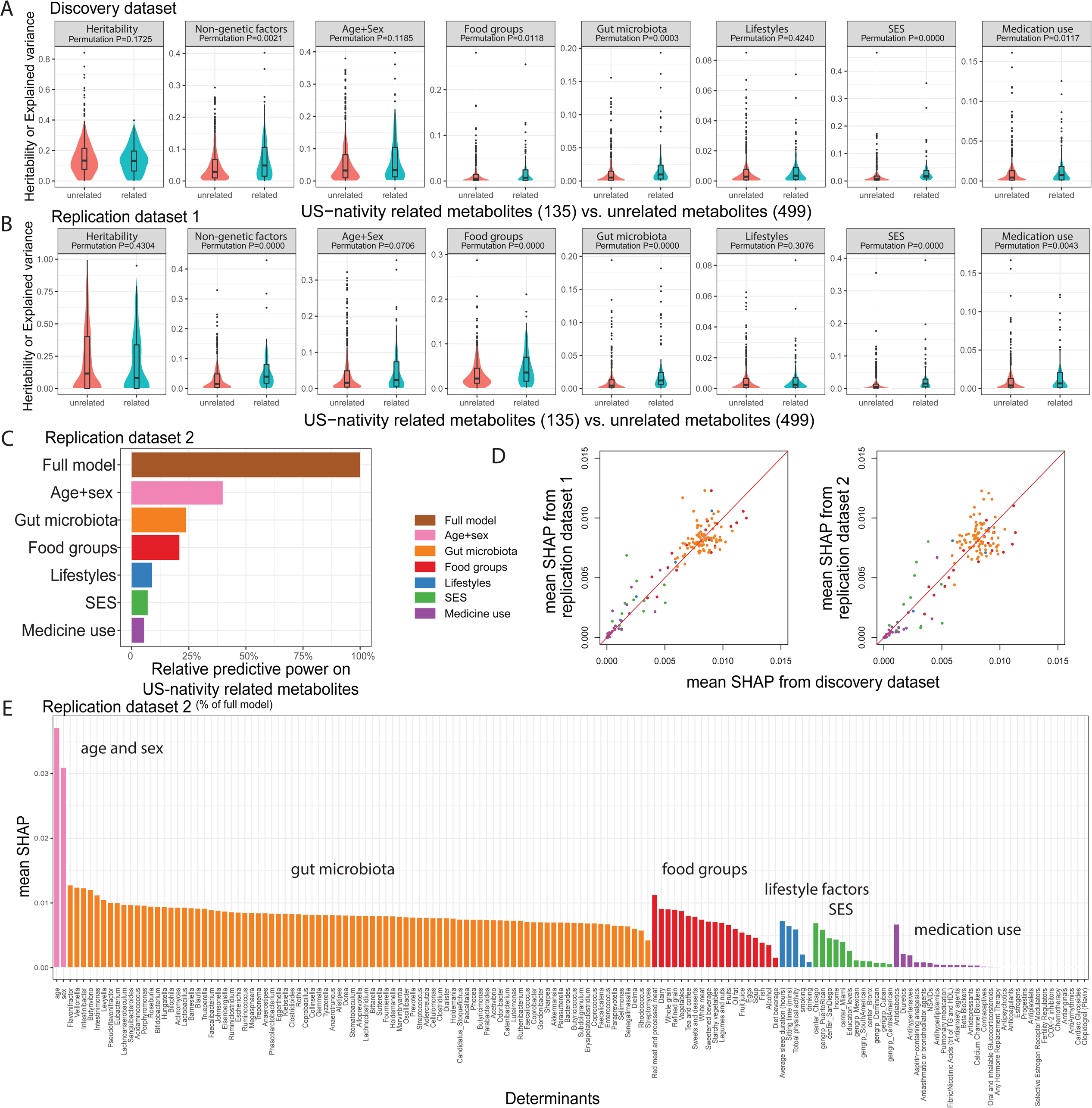
**Determinants of U.S. nativity related metabolomic difference**. (A, B) Boxplot showing the variance of each metabolite explained by each determinant category or SNP-based heritability for 135 U.S. nativity related metabolites (green) and 499 unrelated metabolites (red). Explained variance (EV; R2) for each metabolite was obtained from XGBoost model which used each determinant group to predict metabolite. The difference of explained variance (EV) and heritability between the two metabolite sets were detected using permutation test (1000 times). Fig. a shows results from discovery dataset, and b from replication dataset. **(C)** Barplot showing the relative predictive power of each determinant category on overall 135 U.S. nativity related metabolites in replication dataset 2. The total predictive power of each determinant group is the sum of the EV of the first 100 principal components weighted by the EV of each principal components (Methods). Relative predictive power for each determinant group is the fraction of the total predictive power of the separate model of determinant group compared to the total predictive power of the full model of all non-genetic determinants. **(D)** Consistency of SHAP values of each determinant from discovery dataset, replication dataset, and replication dataset 2. **(E)** Mean SHAP value of each determinant extracted from the full model (Fig. C) in replication dataset 2.

We then examined the strength of association with the overall U.S. nativity related metabolite profile, assessed by the Top 100 principal component of 135 U.S. nativity related metabolites. This was accomplished by a multivariate model which included all non-genetic factors in replication dataset 2 where blood metabolome and gut microbiota data were assessed concurrently. Among these non-genetic factors, age and sex had the highest predictive power (39.4% of the full model), followed by gut microbiota (23.7%) and food groups (20.8%) (**Figure 3C**). To examine the contribution of each individual non-genetic determinant to each U.S. nativity related metabolite, we extracted SHAP values of these non-genetic determinants in the full model on each participant for each metabolite, and then averaged SHAP values across all participants for each individual non-genetic determinant. This process was performed in discovery dataset, replication dataset 1 and replication dataset 2 (**Table S5**). Age, sex, and individual determinants from gut microbiota and food group categories showed higher mean SHAP values compared to other non-genetic factors, and results were consistent across different datasets (**Figure 3D**). **Figure 3E** shows mean SHAP values of each individual determinant on 135 U.S. nativity related metabolites in replication dataset 2.

### Food groups and gut microbes associated with U.S. nativity related metabolites

Since food groups and gut microbiota were major determinants of U.S. nativity related metabolites, we further examined the associations of food groups and gut microbiota with these metabolites. Among 135 U.S. nativity related metabolites, we found 71 metabolites (34 higher in U.S.-born individuals and 37 higher in foreign-born individuals) with EV≥2.5% by food groups and/or gut microbiota (without adjustment for each other) in the replication dataset 2. This included 45 metabolites primarily associated with food groups (EV range, 2.6%-23.3%), 10 metabolites primarily associated with gut microbiota (EV range, 2.8%-18.8%), and 16 metabolites associated with both (EV range, 3.0%-23.0% for food groups; 3.0%-19.0% for gut microbiota) (**Figure 4A**). These results were further confirmed by including both food groups and gut microbiota in the same model (i.e., adjustment for each other) (**Figure 4B**).

**Figure 4.**
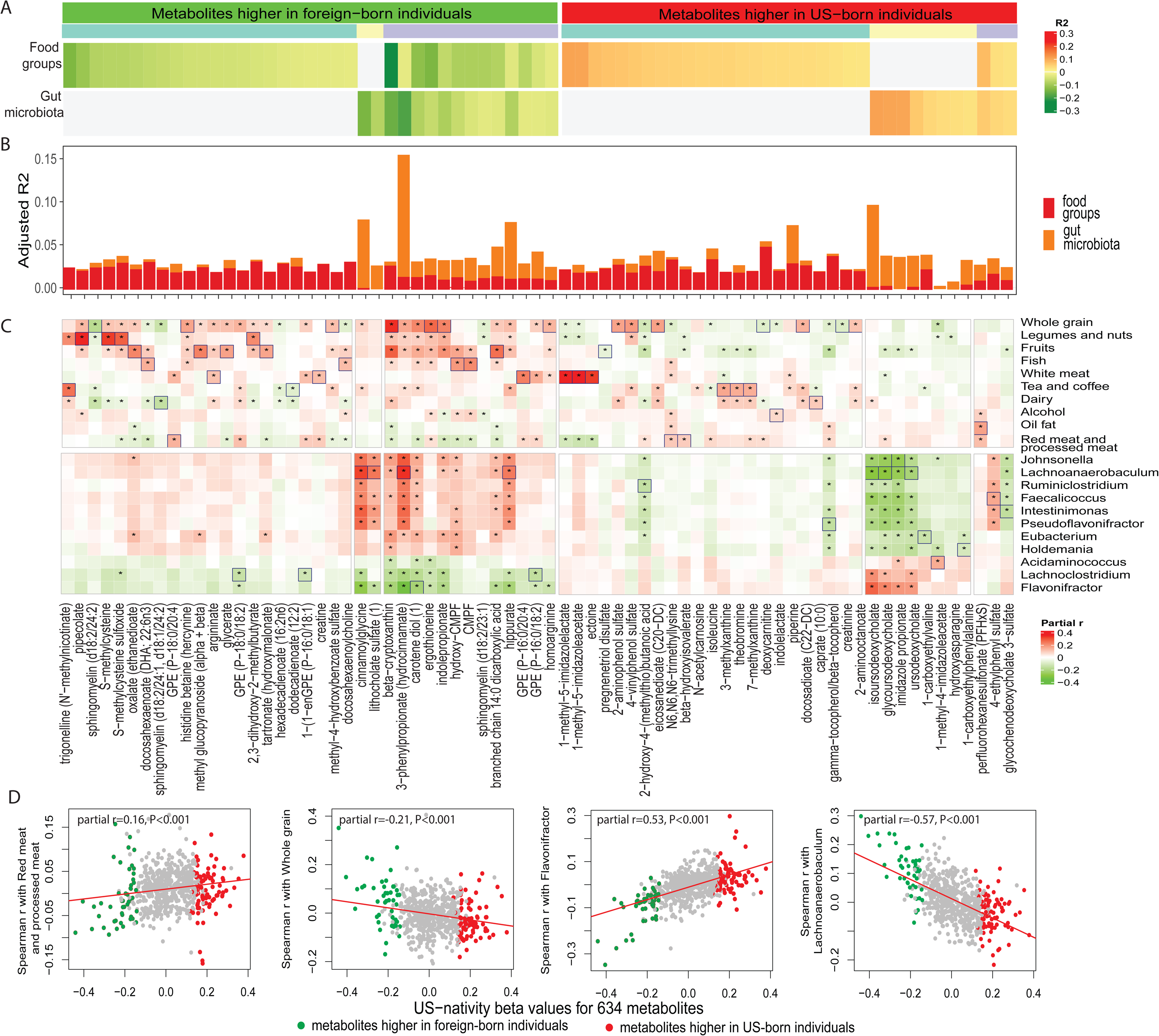
**U.S. nativity related metabolites primarily determined by food or gut microbiota**. (A) Heatmap showing explained variance (EV) of metabolites (columns) which are primarily determined by food or gut microbiota (rows). Yellow gradient colors indicate the EV of metabolites higher in the blood of U.S.-born participants compared to foreign-born participants, green gradient colors indicate the EV of metabolites higher in the blood of foreign-born participants, and gray indicates the EV of metabolites not primarily determined by food or gut microbiota. Metabolites higher among U.S.-born or foreign-born participants are further divided into three groups: those primarily explained by food groups (food-derived metabolites), those primarily explained by gut microbiota (microbiota-derived metabolites), and those explained by both (food and microbiota-derived metabolites). (B) Adjusted variance explained by food groups and gut microbiota (adjusted explained variance; EVadj). The adjusted variance explained by food groups was calculated based on the residuals of linear model fitting metabolite and gut microbiota, and vice versa. (C) Heatmap showing partial Spearman’s correlation coefficient between metabolites and individual food group or microbe. Red indicates positive correlation, green indicates negative correlation, and marked correlations are those statistically significant after FDR adjustment (FDR<0.05). The strongest correlation for each metabolite is highlighted in a boxed cell. Only individual food groups and microbes with at least one significant correlation with metabolites are shown. Columns in Fig a, b, and c are aligned to show the same metabolite. (D) Correlation between the effect size of U.S. nativity on metabolites and the effect size of four selected determinants on metabolites. 71 selected metabolites from A-C are colored in red (metabolites higher in U.S.-born individuals) or green (metabolites higher in foreign-born individuals) among 643 profiled metabolites. Pearson’s correlation coefficients and P-value are shown in each panel.

Metabolites primarily associated with food groups (N=45) are from sub-pathways, including Xanthine Metabolism, Histidine Metabolism, Food Component/Plant, Leucine, Isoleucine and Valine Metabolism, etc. This set was correlated with multiple individual dietary factors but few gut bacterial genera (**Figure 4C**). Metabolites primary associated with gut microbiota (N=10) are from Secondary Bile Acid Metabolism, Histidine Metabolism, etc., and they were correlated with multiple gut bacterial genera but few dietary factors (**Figure 4C**). Metabolites primary associated with both food groups and gut microbiota (N=16) are from Benzoate Metabolism, Fatty Acid, Dicarboxylate metabolism, Plasmalogen, Vitamin A Metabolism, etc., and they were correlated with both multiple dietary factors and gut bacterial genera (**Figure 4C**). In general, metabolites higher in foreign-born individuals were positively correlated with Johnsonella, Lachnoanaerobaculum, Ruminiclostridium, Faecalicoccus, Intestinimonas, Pseudoflavonifractor, Eubacterium and Holdemania, and were inversely correlated with Acidaminococcus, Lachnoclostridium, and Flavonifractor, while metabolites higher in U.S.-born individuals were correlated with these gut bacterial genera in opposite directions (**Figure 4C**; Lachnoanaerobaculum and Flavonifractor as examples in **Figure 4D**). Significant correlations were observed for most metabolites primarily determined by gut microbiota and those determined by both gut microbiota and food groups. Similarly, metabolites higher in foreign-born individuals were positively correlated with healthy plant-based food intake (e.g., whole grain, legumes and nuts, fruits) and inversely correlated with unhealthy food intake (e.g., red meat and proceed meet, oil fat), while metabolites higher in U.S.-born individuals were also correlated with these dietary factors in opposite directions (**Figure 4C**; whole grain and red meat/processed meat as examples in **Figure 4D**).

### Association between U.S. nativity related metabolites and health outcomes

We conducted a thorough examination of various clinical traits (**Table S6**) and disease conditions among US Hispanics (**Table S7**) to investigate their associations with U.S. nativity related metabolites in the discovery dataset and replication dataset 1, separately. We used 135

U.S. nativity related metabolites to predict each clinical trait and calculated EV for each clinical trait by these metabolites in the discovery dataset and replication dataset 1. The estimated EV of clinical traits by these metabolites were highly consistent between two datasets (**Figure 5A**). The top clinical traits with EV>50% by these metabolites were related to kidney function (Cystatin C, creatinine, eGFR), glycemic traits (fasting glucose, HbA1C), liver function (AST) and blood lipids (TC) (**Figure 5A**). We also compared EVs of clinical traits by U.S. nativity related metabolites and EVs by U.S. nativity unrelated metabolites. As compared to U.S. nativity unrelated metabolites, U.S. nativity metabolites showed higher EV for BMI, TC, TG, fasting glucose, and eGFR, but lower EV for AST in both datasets (**Figure 5B-C**).

**Figure 5.**
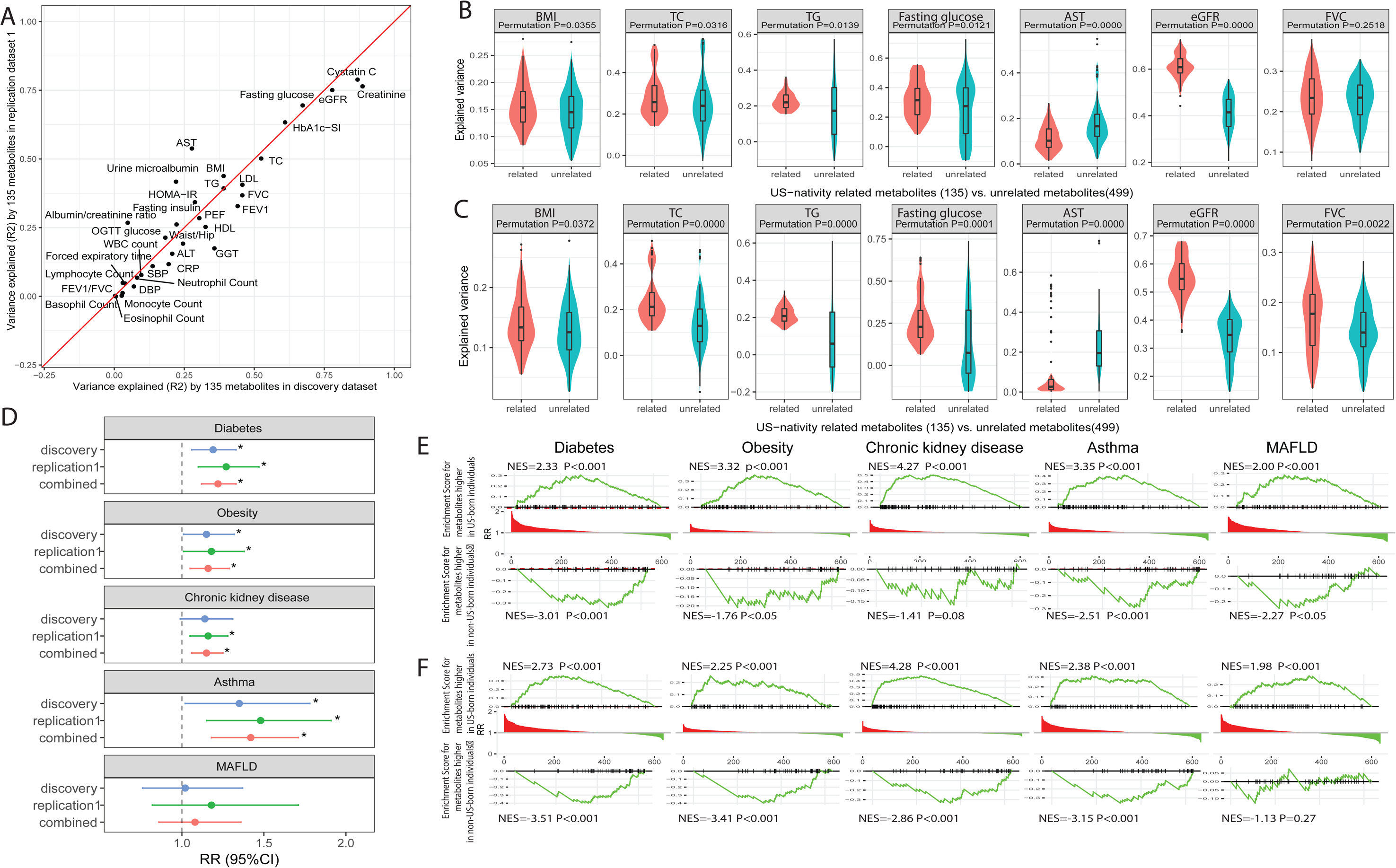
Association between U.S. nativity related metabolites, clinical traits and diseases. (A) Explained variance (EV) of selected clinical traits predicted by 135 U.S. nativity related metabolites in discovery dataset (x-axis) and replication dataset (y-axis). b-c, Comparing EV of selected clinical traits predicted by U.S. nativity related metabolites and that predicted by U.S. nativity unrelated metabolites. To avoid the bias of predictive power due to the different number of metabolites between two metabolite sets, 25 metabolites were randomly selected from 135 U.S. nativity related metabolites and were used to predict each clinical trait. The process was repeated 500 times, generating 500 EV values. The same process were performed on 499 U.S. nativity unrelated metabolites, generating 500 EV values. (B) Boxplot of EV values of U.S. nativity related metabolites (red) and these of U.S. nativity unrelated metabolites (green) from discovery dataset. (C) Boxplot of EV values of U.S. nativity related metabolites (red) and these of U.S. nativity unrelated metabolites (green) from replication dataset. Comparison of these two sets of EV values was carried out using permutation test. (D) Relative risk (RR) and 95% confidence interval (95% CI) of metabolomic score for diabetes, obesity, chronic kidney disease, asthma, and liver diseases in discovery dataset (blue), replication dataset 1 (green), and the combined RR (red). (E-F) Enrichment of U.S. nativity related metabolites in disease-associated metabolites in discovery dataset (fig. e) and replication dataset (fig. f). 634 metabolites are ranked by their association with each disease and aligned along x-axis in each subplot. Metabolites with positive association are on the left, and metabolites with negative associations are on the right. Rug plot along x-axis shows the positions of U.S. nativity related metabolites among the ranked metabolites. Metabolite Set Enrichment Analysis (MSEA) was used to calculate the enrichment score. Normalized enrichment score (NES) and statistic significance are reported at the top. Fig.e and f show the data from discovery dataset and that from replication dataset 1, respectively.

We then examined, during ∼6 years of cohort follow up, the prospective associations of the metabolomic score and risk of diabetes, obesity, chronic kidney disease (CKD), asthma, and MAFLD. The metabolomic score was significantly associated with higher risk of diabetes (Relative risk [RR]=1.22, 95% confidence interval [95% CI]=[1.12-1.33], per SD increment), obesity (RR [95% CI]=1.16 [1.05-1.29]), CKD (RR [95% CI]=1.15, [1.06-1.25]), and asthma (RR [95% CI]=1.42, [1.18-1.71]), but was not associated with risk of MAFLD (RR [95% CI]=1.08 [0.86-1.36]). To further examine the associations between U.S. nativity-related metabolites and risk of these chronic diseases, we additionally performed metabolite set enrichment analysis of U.S. nativity related metabolites in all metabolites ranked by their associations with incident risk of diseases. U.S. nativity related metabolites were enriched in metabolites associated with risk of diabetes, obesity, CKD and asthma, but not in those associated with risk of liver disease, in both datasets (**Figure 5E-F**).

## Discussion

Over the last few decades, epidemiological research on U.S. immigrants has drawn significant attention for two primary reasons: firstly, health disparities among immigrants contribute to a cascade of disease burdens, impacting the overall public health of populations [2, 3, 28]; and secondly, immigrants often experience a range of environmental and lifestyle-related risk factors, not only affecting their own health but also providing insights into causal factors affecting others in a society marked by chronic disease escalation due to the influence of an increasingly industrialized lifestyle [16, 29, 30]. Such research offers crucial insights into the broader impact of lifestyle on health and disease susceptibility among diverse populations. Hispanics/Latinos are a large ethnic minority population in the U.S., with a large heterogeneity from immigration status, cultural, socioeconomic, and genetic perspectives [31], which provides a unique opportunity to study the effect of U.S. lifestyle on health. A thorough assessment of the relationship between risk factors, the disease-informative blood metabolome, and health outcomes could be a critical component of disease prevention.

In this study, we observed that a substantial proportion (21.1%) of measured blood metabolites in a broad commercially available panel could collectively be used to distinguish U.S.-born and foreign-born Hispanics/Latinos in the U.S. The metabolites associated with U.S. nativity also correlated to other immigration-related variables. These observations underscore the profound impact of U.S. lifestyle on blood metabolome among immigrants to the U.S. Previous studies have shown that a broad spectrum of factors, including host genetics, diet, gut microbiome, clinical parameters, lifestyle and anthropometric measurements, can influence the blood metabolome [32]. This suggests that certain risk factors inherent in U.S. lifestyle contribute to a significant shift in the blood metabolome observed among Hispanics/Latinos in the U.S.

After extensively examining the associations of potential risk factors with the metabolomic difference between the two nativity groups and their associations with individual U.S. nativity related metabolites, we found that age and sex did not contribute to the metabolic difference but were the two prominent contributors to U.S. nativity related metabolites, although they were adjusted as covariates in the multiple linear regression model to identify U.S. nativity related metabolites. The influence of age and sex on blood metabolites have been well established in numerous studies [33–36], and our study further underlines their significance as crucial covariates in blood metabolomics studies. Despite the anticipation that genetic factor would be irrelevant to the metabolomic difference between U.S.-born and foreign-born Hispanics, we noted some metabolites with high heritability (h^2^_snp_>0.30), aligning with previous findings [7], and some of the heritable metabolites were related to U.S. nativity. While the full genetic effect on U.S. nativity related metabolites is beyond our current research scope, it is worthy to further explore the metabolites determined by both genetics and environmental factors in the future studies, which could reveal certain genotypes which are more sensitive to the impacts of a U.S. lifestyle.

We discovered that, among a range of modifiable factors that were examined, gut microbiota and food groups represent the primary potential determinants of the metabolomic difference between the two U.S. nativity groups. Recent studies on blood metabolites have consistently revealed that food and gut microbiota are important sources of blood metabolites [7, 8, 11, 37]. Our study, going beyond confirming the important roles of diet and the gut microbiome in shaping the metabolome, focuses on prioritizing specific factors within these two categories using an interpretable machine learning based analysis approach. Upon examining feature importance among all available food groups, we identified that red and processed meat, dairy, whole grain, vegetables, tea and coffee, and sweets and desserts were top determinants of overall U.S. nativity related metabolites. The observation of increased consumption of red and processed meat, dairy, refined grain, and sweets and desserts, along with reduced intake of whole grain and vegetables among U.S.-born Hispanics/Latinos, mirrors their dietary shift in a U.S. lifestyle. While many prior studies have reported the association between these food types and blood metabolites [38–41], our study provides a deeper understanding of the relationship between these food groups and blood metabolites within context of U.S. nativity, which might offer direction for nutritional studies aimed at understanding chronic diseases related to U.S. acculturation.

Among gut microbiota, we identified *Flavonifractor, Veillonella, Intestinibacter, and Butyrivibrio* as the top microbes associated with U.S. nativity related metabolites. *Flavonifractor* exhibited higher abundance among U.S.-born Hispanics/Latinos, while the others showed a lower abundance. *Flavonifractor*, an intestinal microbiota which enhances oxidative stress, has been associated with bipolar disorders [42] and affective disorders [43]. Despite its unknown metabolic function, its representative species, *Flavonifractor plautii*, is inversely associated with consumption of flavonoids, which are a group of polyphenolic dietary compounds found in many different plant-based foods [44]. *Veillonella* plays a significant role in using lactic acid for primary carbohydrate metabolism in the human small intestine [45]. *Intestinibacter*, although known as a protective microbe for constipation, lacks clear insight into its metabolic functions [46]. *Butyrivibrio*, extensively studied for its Carbohydrate-Active enZYmes (CAZymes) that break down plant-derived polymers, also produces health-promoting compounds like conjugated linoleic acid and vitamins [47]. Collectively, these findings suggest that U.S.-born Hispanics/Latinos have less favorable gut microbiome composition as compared to their counterparts, which may contribute to metabolomic shift.

As gut microbiota and food groups were found to be two important sources of blood metabolites in previous studies and in our study, a noteworthy aspect of our study is the ability to examine their synergistic or independent impact on U.S. nativity related metabolites. Focusing on the top metabolites determined by either of these two groups of factors, we discerned a distinct trend regarding their associations with specific food groups and microbes. Although our current cross-sectional study cannot establish causal relationships between these sources, it revealed that certain U.S. nativity related metabolites were influenced by both gut microbiota and food groups, underscoring the significant interplay between dietary choices and gut microbiota in shaping blood metabolites [7, 8, 38].

Finally, we discovered that U.S. nativity related metabolites were collectively associated with representative clinical traits reflecting blood glycemic and lipid profiles, liver function, kidney function, and lung function, to a greater extent than metabolites unrelated to U.S. nativity. Additionally, U.S. nativity related metabolites were collectively overlapped with these metabolites associated with representative common diseases that are prevalent among Hispanics/Latinos in the U.S. Hispanics/Latinos in the U.S. have a high burden of multiple chronic diseases [31, 48, 49], and prior studies have attempted to pinpoint one or multiple causal factors [13, 50, 51]. However, our study suggests that a systemic metabolomic shift in metabolic patterns, resulting from the adoption of U.S. lifestyle due to multiple risk factors, significantly contributes to this multifaceted disease burden.

Our study has some limitations that should be considered. First, the analysis focused on Hispanics/Latinos in the U.S., which may limit the generalizability of the findings to other U.S. immigrant groups, or people of Latin American birth living in the other global regions. Second, the study relied on cross-sectional data, which limits the ability to establish causal relationships between determinants, metabolomic differences, and health outcomes. Third, longitudinal studies would be valuable to better understand the dynamic changes in the metabolome over time with immigration to the U.S.

In the current study, we used comprehensive profiling of the blood metabolome, gut microbiota, genotypic data, environmental factors, and health outcomes for U.S.-born and foreign-born Hispanics/Latinos in the U.S. to investigate the relationship among risk factors, blood metabolites related or unrelated to U.S. nativity, and multiple clinical traits and chronic diseases. The difference in metabolomic profiles by U.S. nativity determined by multiple risk factors particularly among food groups and gut microbiota, shed light on the impact of U.S. acculturation on health and provides vital clues for prevention of chronic disease burden in immigrant communities. Understanding the association between metabolomic variations, risk factors, and disease outcome could pave the way for tailored interventions aiming to mitigate the health risks associated with U.S. lifestyle during acculturation process.

## Data Availability

All data produced in the present study are available upon reasonable request to the authors

## Acknowledgements

The Hispanic Community Health Study/Study of Latinos is a collaborative study supported by contracts from the National Heart, Lung, and Blood Institute (NHLBI) to the University of North Carolina (HHSN268201300001I / N01-HC-65233), University of Miami (HHSN268201300004I / N01-HC-65234), Albert Einstein College of Medicine (HHSN268201300002I / N01-HC-65235), University of Illinois at Chicago (HHSN268201300003I / N01-HC-65236 Northwestern Univ), and San Diego State University (HHSN268201300005I / N01-HC-65237). The following Institutes/Centers/Offices have contributed to HCHS/SOL through a transfer of funds to the NHLBI: National Institute on Minority Health and Health Disparities, National Institute on Deafness and Other Communication Disorders, National Institute of Dental and Craniofacial Research, National Institute of Diabetes and Digestive and Kidney Diseases (NIDDK), National Institute of Neurological Disorders and Stroke, and NIH Institution-Office of Dietary Supplements.

This work is supported by the National Institute of Environmental Health Sciences (R01ES030994) and the National Institute of Diabetes and Digestive and Kidney Diseases (R01DK119268). Other funding sources for this study include R01HL060712, R01HL140976, R01HL105756, and R01HL136266 from the NHLBI; and R01DK120870, the New York Regional Center for Diabetes Translation Research (P30 DK111022), Diabetes Research Center (DRC) grant DK063491 to the Southern California Diabetes Endocrinology Research Center, and UM1DK078616 from the NIDDK; and the National Center for Advancing Translational Sciences, CTSI grant UL1TR00188. This research has been conducted using the UK Biobank Resource under Application Number 56483.

## Supplemental Information

Supplemental methods.doc

Supplemental tables.xlsx

Table S1 Baseline characteristics of participants in three datasets by U.S. nativity

Table S2 Baseline characteristics of age-matched participants by U.S. nativity

Table S3. Association between U.S. nativity and metabolites in age-matched discovery dataset and replication dataset 1

Table S4. Metabolite heritability and Metabolite variance explained by each determinant category from XGBoost model in discovery dataset and replication dataset 1

Table S5. Mean SHAP values of each determinant in predication model of metabolites from XGBoost model in three datasets

Table S6. Variance of clinical traits explained by U.S. nativity-related metabolites in discovery dataset and replication dataset 1

Table S7. Association between metabolites and incidence of selected diseases in discovery dataset and replication dataset 1

