## Supplemental methods for "Blood metabolomic shift links diet and gut microbiota to multiple health outcomes among Hispanic/Latino immigrants in the U.S."

**Author affiliations**

### Study cohort

The HCHS/SOL is a prospective study of Hispanic/Latino populations in the U.S. From 2008 to 2011, 16 415 adults aged 18–74 years, self-identified as having Cuban, Dominican, Puerto Rican, Mexican, Central American or South American heritage, were recruited from a random sample of households in four communities (Bronx, NY; Chicago, IL; Miami, FL; and San Diego, CA). Participants were recruited by using a 2-stage probability sample design. A comprehensive battery of interviews and a clinical assessment with fasting blood draw were conducted by trained and certified staff at in-person clinic visits from 2008 to 2011 (visit 1). The second visit (visit 2) period started in October 2014 and concluded in December 2017. A total of 11,623 cohort members were re-examined in visit 2 to collect data predictive of cardiopulmonary outcomes and the onset of diabetes. The study protocol was approved by the institutional review boards of all collaborating institutions, and written informed consent was obtained for all participants.

The data for the entire cohort were divided into discovery dataset and validation dataset based on the batch of blood metabolomic profiling while taking into account the visit at which blood sample were collected. The first batch of 3,941 blood samples for metabolomic profiling were collected at the baseline visit. The second batch of blood samples from 3,178 participants were profiled, of which 2,368 samples were collected at baseline visit and 810 samples at six-year follow-up visit. Thus, we divided all participants into three datasets: discovery dataset (N=3,941. Blood samples from baseline visit; metabolome was profiled at the first batch), replication dataset 1 (N=2,368. Blood samples from baseline visit; metabolome was profiled at the second batch), and replication dataset 2 (N=810. Blood samples from follow-up visit; metabolome was profiled at the second batch). This division aimed to address the potential batch effects in metabolomic profiling caused by profiling batch and this visit.

Discovery dataset and replication dataset 1 served to identify and confirm U.S.-nativity related blood metabolites. To minimize the impact of age on blood metabolome, we conducted age matching at 1:1 ratio within discovery dataset and 1:2 ratio within replication dataset, according to the proportion of U.S.-born and foreign-born participants. The procedure aimed to create sub-datasets for specific analyses.

All stools for shotgun metagenomic sequencing were collected at the follow-up visit, and microbial composition for all individuals was profiled at the same batch. There was not six-year gap between blood metabolome and stool metagenome in the replication dataset 2. As such, we used the discovery dataset and replication dataset to characterize U.S. nativity related metabolomic difference and identify potential determinants, while replication dataset 2 was specially used to compare the contribution of determent group to metabolomic difference and prioritize each individual determinant.

### Immigration/acculturation-related variables

We used U.S. nativity as the main immigration variable to examine the influence of immigration on blood metabolome. Other five immigration/acculturation-related variables were also collected or calculated to validate U.S. nativity related metabolomic change.

Immigration age: age of immigration among participants who were not born in US mainland.

Years lived in the U.S.: years lived in the U.S. mainland for foreign-born Hispanics.

Dietary acculturation score is based on the U.S.ual food consumption, with five possible values: 1 = Mainly Hispanic/Latino foods, 2 = Mostly Hispanic/Latino foods and some American food, 3 = Equal amounts of both Hispanic/Latino and American foods, 4 = Mostly American foods and some Hispanic/Latino foods, and 5 = Mainly American foods.

Language acculturation score is based on the language spoken at home, with five possible values: 1 = Only Spanish, 2 = More Spanish than English, 3 = Both equally, 4 = More English than Spanish, and 5 = Only English.

Social acculturation score is the average of four 5-point questions: (1) Your close friends are? (2) You prefer going to social gatherings/parties at which the people are? The people you visit or who visit you are? (3) If you could choose your children’s friends, (4) you would want them to be?. Each question has five values: 1 = All Hispanic/Latino, 2 = More Hispanic/Latino than non-Hispanic/non-Latino, 3 = About half and half, 4 = More non-Hispanic/non-Latino than Hispanic/Latino, and 5 = All non-Hispanic/non-Latino. The average of the 4 questions indicates the degree of social acculturation.

### Metabolomic profiling of blood samples

Blood metabolomic profiling was performed using an untargeted liquid chromatography-mass spectrometry (LC-MS) based protocol on discovery HD4 platform at Metabolon (Durham, North Carolina, USA). Participants were asked to fast for ≥8 h before the examination, consume only water and necessary medications, and to refrain from smoking or physical activity before undergoing the fasting examination procedures. Detailed procedures have been described elsewhere [1]. After removing unknown metabolites and the metabolites which were undetected in greater than 20% samples, 634 metabolites were finally obtained for the following analyses in the current study. Annotation of these metabolites included their super-pathway and sub-pathway, which were provided by Metabolon Inc. All these metabolites were transformed using Inverse normal transformation.

### Metagenomic profiling of Stool samples

Metagenomics Sequencing was performed on DNA extracted from fecal samples collected by FTA card using a novel shallow-coverage method of shotgun sequencing-based Illumina NovaSeq platforms. Microbiome bioinformatics analyses, taxonomic assignment, and functional components identification were performed using the SHOGUN pipeline as described previously [2]. Genus level data was used in the current analyses to reduce the number of microbe features for two considerations: (1) Species from the same genus have similar characteristics and functions; (2) A large number of features will lead to an inflation of proportion of the variance for a dependent variable that's explained by independent variables in the regression machine learning model compared to the model including few variables. Metagenomic data was normalized and transformed using Centered Log-Ratio method (CRL). After removing these genus which were prevalent in less than 20% samples or whose average read counts across samples were less than 10, 87 genus were included in the following analyses.

### Identification of US-related metabolites and Derivation of metabolomic score

Multiple linear regression model with each metabolite as dependent variable was used to identify different metabolites between U.S.-born participants and foreign-born participants while controlling for age, sex, visit center, Hispanic background, and five principal components of population structure. As metabolites were normalized, the beta values from the Multiple linear regression model were used to measure the effect size of effect of U.S. nativity on each metabolite. These metabolites which were identified in the matched discovery dataset (P<0.05 after False Discovery Rate [FDR] correction) and confirmed in the matched replication dataset 1 (P<0.05) were defined as U.S. nativity related metabolites. The rest metabolites were considered as U.S. nativity unrelated metabolites.

Least Absolute Shrinkage and Selection Operator (LASSO) was applied to predict U.S. nativity status (U.S.-born as 1 and foreign-born as 0) with U.S. nativity related metabolites using glmnet package, from which the predictive value was generated and used as metabolomic score. The training and testing of LASSO model were performed in different datasets. Tenfold cross-validation in training dataset was used to select the optimal tuning parameter for LASSO. The predictive performance of metabolomic score in discriminating U.S.-born and foreign-born participant was measured by calculating Area Under the Receiver Operating Characteristic (AUROC).

Metabolomic score represented the summary of metabolomic difference between U.S.-born participants and foreign-born participants. Its association with other immigration-related variables was further examined. Meanwhile, years lived in the U.S. were discretized to examine the dose-response relationship with metabolomic score.

### Determinant categories

The ‘genetics’ includes all genotypic SNPs.

The ‘age+sex’ includes age and sex.

The ‘food groups’ includes 19 food groups aggregated based on Food Group Serving at the Daily Totals Level with 167 food types. There are Red meat and processed meat, Dairy, Whole grain, Refined grain, Vegetables, Tea and coffee, Sweets and desserts, White meat, Sweetened beverage, Starchy vegetables, Legumes and nuts, Fruits, Oil fat, Fruit juice, Eggs, Soup, Fish, Alcohol and Diet beverage.

The ‘gut microbiota’ includes 87 microbes at the genus level as mentioned above.

The ‘lifestyle’ includes smoking status (never, former, and current), years of cigarette smoking, drinking status (never, former, and current), drinks per week, total physical activity (from Global Physical Activity Questionnaire), sedentary time (sitting or reclining time on a typical day, mins), and weekday sleep duration (hours).

The ‘Socioeconomic status (SES)’ includes Income, Education level, visit center, and Hispanic background. Four visit centers (Chicago, Bronx, San Diego, and Miami) and five Hispanic background (Puerto Rican, Mexican, South American, Dominican, Cuban, and Central American) were processed as binary variable using one hot encoding method.

The ‘medication use’ group includes 20 drugs: Antidiabetics, Diuretics, Antihypertensives, Aspirin-containing analgesics, Antiasthmatic or bronchodilator agents, NSAIDs, Antihyperlipidemics, Pulmonary medication, Fibric/Nicotinic Acids, Antianxiety agents, Beta Blockers, Antidepressants, Calcium Channel Blockers, Contraceptives, Oral and inhalable Glucocorticorsteroids, Any Hormone Replacement Therapy, Antipsychotics, Anticoagulants, Estrogens, Progestins, Antiplatelets, Selective Estrogen Receptor Modulators, Fertility Regulators, COX-2 inhibitors, Chemotherapy, Antianginals, AntiArrhythmics, Cardiac Glycosides, Clopidogrel.

### Association between determinant category and each metabolite

Explained variance (EV) was calculated to measure the association between each metabolite and each determinant category using eXtreme Gradient Boosting (XGBoost) implemented in xgboost package. XGBoost is an extension of the GB classifier, which also focuses on speed and performance. XGBoost includes the regularized learning that helps smooth the final learned weight to avoid overfitting. To estimate the EV of each metabolite, determinants of each category were used as input for XGBoost model, and each metabolite level as output. All XGBoost models were trained using 80% samples in the dataset and testing in 20% samples. Permutation test was used to compare the EV values of U.S. nativity related metabolites and unrelated metabolites for each determinant category.

Adjusted EV (EV_adj_) was additionally calculated for food groups and gut microbiota. To calculate EV_adj_ for food groups, the first 30 PCs from Principal Component Analysis (PCA) of gut microbiota were used in a Multiple linear regression model with metabolite level as output. Residuals from the regression were considered as metabolite level adjusted by gut microbiota and were then used as output for XGBoost with food groups as input. The EV from the XGBoost model was considered as EV_adj_ for food groups with adjustment for gut microbiota. Similarly, EV_adj_ for gut microbiota with adjustment for food groups was calculated.

### Genotyping and SNP-based heritability of metabolites

Genotyping and imputation protocols have been described previously [3]. Genotyping was performed by Illumina Microarray Services with a custom array. Genotype data were cleaned, and quality checked centrally at Illumina Microarray Services, LA Biomed, and the University of Washington, which heads the HCHS/SOL Genetic Analysis Center. There were >25.5 million imputed genotypes, for a total of 27.7 million tested SNPs. We excluded variants with low minor allele counts (<30) and low imputation quality (<0.3) from all results shown. Detailed procedures have been described elsewhere [4].

Heritability for each metabolite were estimated via a mixed model implement in GENESIS package using the variance explained by the kinship matrix, representing the variance explained by additive effects of common genetic variants. Heritability was estimated in participant after excluding >3rd-degree relatives estimated via the kinship coefficient.

### Feature importance of determinants

To measure the feature importance of each determinant and find specific associations between features and metabolite levels, Shapley additive explanations (SHAP) values were calculated based a full model including all non-genetic determinants. SHAP value is a recently introduced framework for interpreting predictions, which assigns each feature the importance value for a particular prediction (sample). In brief, for a specific prediction, the SHAP value of a feature is defined as the change in the expected value of the output of the model when this feature is observed versus when it is missing. The mean SHAP values for each determinant were finally calculated as the averaged absolute SHAP value for this determinant across all samples, which reflected the mean effect of each feature on the predictions and served as a feature importance measure. SHAP values for XGBoost were calculated using SHAPforxgboost package.

### Predictive power of determinants on overall U.S. nativity related metabolites

Predictive power on each metabolite was estimated for each determinant category using XGBoost, except genetic factors which used heritability instead of predictive power. To estimate the predictive power of different determinant category (separated model) and all determinants (full model), we first applied PCA over all U.S. nativity related metabolites data to get the first 100 PCs which constitute more than 98% of the total variance in the data. XGBoost was used to predict the PCs on the basis of the determinant categories independently. EV of each PCs were calculated separately and then combined as predictive power for each determinant category:

$$\frac{\Sigma_{ⅈ=1}^{n}EV_{i}\times PC_{i}}{\Sigma_{ⅈ=i}^{n}PC_{i}}$$

where EV_i_ is the EV that XGBoost recovered for the i^th^ PC. PC_i_ is the fraction of variance that the i^th^ PC explains out of the overall variation of 100 PCs. n is the total number of PCs.

A full model which included all non-genetic determinants was built to predictive power for all non-genetic determinants. Relative predictive power was the percentage of predictive power of each category model in the full-model predictive power.

### Ascertainment of health outcomes

The investigated clinical traits include continuous variables from cardiometabolic traits, chronic inflammation marker, blood cell counts, liver function indices, kidney function indices, and lung function indices.

Cardiometabolic traits had Body Mass Index (BMI), Waist/hip ratio, Triglycerides (TG), Total cholesterol (TC), LDL-cholesterol (LDL-C), HDL-cholesterol (HDL-C), Fasting insulin, Fasting glucose, 2h Oral glucose tolerance tests (2h-OGTT) glucose, Glycosylated Hemoglobin in SI units (HbA1c-SI), Homeostatic Model Assessment for Insulin Resistance (HOMA-IR), Systolic blood pressure (SBP), and Diastolic blood pressure (DBP).

Chronic inflammation marker was C-reactive protein (CRP).

Blood cell counts had White blood cell (WBC) count, Neutrophil Count, Lymphocyte Count, Monocyte Count, Eosinophil Count, and Basophil Count.

Live function indices had Alanine transaminase (ALT), Aspartate transaminase (AST), and Gamma-glutamyl transferase (GGT).

Kidney function indices had Estimated glomerular filtration rate (eGFR), Cystatin C, Creatinine, Urine microalbumin, and Albumin/creatinine ratio (ACR).

Lung function indices had Forced vital capacity (FVC), Forced expiratory volume in 1 second (FEV1), FEV1/FVC Ratio, Peak expiratory flow (PEF), and Forced expiratory time.

For comparison among different traits, all clinical traits were standardized if their variance was analyzed.

Five representative diseases were investigated in the study, including type 2 diabetes (T2D), obesity, chronic kidney disease (CKD), asthma, and chronic liver disease.

Diabetes: we examined two definitions of incident diabetes in order to enhance comparisons of our results with other studies that have used various diabetes definitions. The first definition was based on three criteria: 1) self-reported diagnosis of diabetes, 2) self-reported use of diabetic medication, or 3) laboratory-tested fasting plasma glucose > 126 mg/dl, OGTT glucose > 200 mg/dl, or HbA1c> 6.5%.

Obesity was defined as BMI ≥30 kg/m2.

CKD was defined as eGFR <60 ml/min/1.73m2 or ACR ≥30 mg/g.

Asthma: Current asthma was defined by a positive answer to all following questions: “Have you ever had asthma?”, “Was it diagnosed by a doctor or other health care professional?”, and “Do you still have it?” For the analysis of current asthma, control subjects were those responding “No” to any of these questions. Current asthma symptoms were independent of a physician’s diagnosis of asthma and were defined by a positive answer to either of these questions: “In the last 12 months, have you had wheezing or whistling in your chest at any time?” or “In the last 12 months, have you been awakened from sleep either by coughing (apart from a cough associated with a cold or chest infection) or by shortness of breath or a feeling of tightness in your chest?” For the analysis of current asthma symptoms, control subjects were those answering “No” to both questions.

Metabolic dysfunction-associated fatty liver disease (MAFLD): we defined suspected MAFLD, which is renamed from Non-alcoholic fatty liver disease (NAFLD), using serum aspartate aminotransferase (AST) and alanine aminotransferase (ALT). Thus, NAFLD was suspected if AST was greater than 37 U/L or ALT greater than 40 U/L in men, and if AST or ALT was greater than 31 U/L in women.
